# Shared epigenetic regulation acting on neuroimmune pathways contributes to the comorbidity between generalized anxiety disorder and COVID-19

**DOI:** 10.64898/2026.06.03.26354830

**Authors:** Sefayet Karaca, Brenda Cabrera-Mendoza, Jun He, Dan Qiu, David Davtian, AnnMarie Lacobelle, Yaira Z. Nunez, John H. Krystal, Robert H. Pietrzak, Joel Gelernter, Renato Polimanti

## Abstract

**Background:** The biological mechanisms linking generalized anxiety disorder (GAD) and COVID-19 remain poorly understood, despite substantial evidence of their comorbidity. To address this gap, we examined genetic and epigenetic factors underlying their co-occurrence.

**Methods:** In a multi-ancestry sample of 893 participants, we conducted genome-wide and epigenome-wide analyses of GAD and COVID-19 severity. Integrating large-scale genome-wide datasets and information regarding methylation quantitative trait loci, complementary analytic approaches were used to identify regional methylation patterns, assess genetically regulated DNA methylation in blood and brain tissue, and evaluate causal loci shared between GAD and COVID-19.

**Results:** GAD was associated with epigenome-wide significant variation in loci involved in chromatin regulation and synaptic signaling. Conversely, COVID-19-related epigenetic signals were enriched in immune-inflammatory and host-response pathways. Mild COVID-19 was epigenetically related to endothelial-inflammatory signals, while severe COVID-19 was linked to epigenetic changes implicated in myeloid and thrombo-inflammatory pathways. Epigenetic signals shared between GAD and COVID-19 implicated processes related to stress adaptation and tissue homeostasis. Genetically informed analyses identified 60 shared loci, including *MAPT*, *ZFP57*, and *FBXL18*, indicating pleiotropy between GAD and COVID-19 in genetically regulated DNA methylation variation. Brain-specific analyses further highlighted convergence in additional loci (i.e., *MICB* and *HLA-DPB1*), suggesting neuroimmune mechanisms underlying GAD-COVID-19 shared methylation patterns.

**Conclusions:** These findings support that GAD and COVID-19 share epigenetic and genetic architecture involving pathways related to vascular integrity, immune function, and cellular adaptation, highlighting a potential neuroimmune basis for their co-occurrence.

## Background

DNA methylation is a key molecular mechanism linking genetic susceptibility and environmental exposures to downstream regulatory processes. Epigenome-wide association studies (EWAS) of anxiety-related phenotypes have identified differential methylation in genes, implicating pathways related to immune regulation, neurogenesis, and stress response [1, 2]. A recent large-scale EWAS of generalized anxiety disorder (GAD) in 43,504 ancestrally diverse participants identified multiple CpG sites reflecting both genetic and environmental influences [3]. The dynamic and environmentally responsive nature of GAD [4] highlights the need for integrative approaches that can distinguish genetically regulated from environmentally responsive epigenetic signatures.

Beyond psychosocial stressors, systemic biological factors have also been linked to increased psychiatric risk [5, 6]. In this context, COVID-19 infection can be particularly relevant given its association with epigenetic remodeling [7–9] and elevated psychiatric outcomes [10]. Epigenome-wide studies have reported widespread methylation alterations following COVID-19 infection, including findings from an early post-infection period [11] and persistent changes during longer-term post-acute follow-up [12, 13]. Furthermore, evolving neuropsychiatric symptoms have also been reported up to 2–3 years after COVID-19 infection [14]. Specifically, inflammation has been shown to induce neurobiological alterations that may increase vulnerability to anxiety disorders [15]. These findings suggest that COVID-19-associated biological stress may induce methylation changes in pathways implicated in anxiety and depression pathogenesis. Overall, COVID-19 provides a particularly relevant model given its psychiatric sequelae and pronounced immune-inflammatory burden [10, 16, 17], making it a useful context for examining shared epigenetic vulnerability underlying infection-related psychiatric outcomes. Despite growing evidence of COVID-19-associated methylation changes at immune and inflammatory loci [18] and their relationship to neuropsychiatric symptoms yet their molecular basis remains poorly understood. To fill this gap, the present study investigated the epigenetic links between COVID-19 and GAD using a multi-omics integrative framework. Specifically, we conducted epigenome-wide and genetically informed analyses in a multi-ancestry cohort to investigate convergent and divergent epigenetic patterns between GAD and COVID-19, and to assess the contribution of genetically regulated mechanisms across blood and brain tissue. Understanding shared mechanisms could elucidate biological pathways linking systemic inflammation to anxiety disorders and guide development of targeted interventions. Additionally, our findings expanded the knowledge of GAD epigenetics, which remains less characterized [3, 19] than that of several other psychiatric disorders [2, 20, 21].

## Methods

This study was conducted under protocol #2000033404 approved by the Human Research Protection Program of the Yale School of Medicine, New Haven, CT, USA. Written informed consent was obtained from all participants in accordance with the Declaration of Helsinki.

### Study Cohort

The study was conducted in a cohort recruited specifically to assess the comorbidity between internalizing disorders and COVID-19. GAD was assessed using the GAD-7 anxiety scale [22]. COVID-19 severity was determined based on self-reported symptoms assessed using the Coronavirus Health Impact Survey [23]. COVID-19 mild symptoms were defined by the presence of fever, cough, sore throat, fatigue, loss of taste or smell, or eye infection, without shortness of breath. Moderate COVID-19 severity was defined by the presence of shortness of breath among reported COVID-19 symptoms. Genetic ancestry was inferred by applying a “k-nearest neighbors” algorithm to top 10 principal components (PC), using 1000 Genomes Project populations as reference panel [24].

### DNA methylation profiling

Genomic DNA (500 ng) was bisulfite-converted using the EZ-96 MagPrep Kit (Zymo Research) according to the manufacturer’s protocol. Following bisulfite conversion, 4 µL of DNA per reaction underwent whole-genome amplification, enzymatic fragmentation, and purification. DNA methylation levels were quantified using the Illumina Infinium MethylationEPIC v2.0 BeadChip (930K). Fluorescent signals were scanned with the Illumina iScan System, and raw intensity data were processed in GenomeStudio to obtain β-values for each CpG site.

### Epigenome-Wide Association Study (EWAS)

DNA methylation data were used for downstream association analyses. We performed ancestry-stratified and cross-ancestry EWAS using the Psychiatric Genomics Consortium pipeline [25, 26]. Standard quality control (QC) procedures were applied and samples failing Illumina control metrics, outliers, and duplicates were excluded. For methylation-specific QC, we applied background correction and noob normalization using the minfi R package [27]. Probes with detection p> 0.01 and CpGs with >10% missing data were removed. No samples were excluded based on signal intensity (<50% of median or <2000 units) criteria. Cross-reactive probes were filtered out, and batch effects were adjusted for technical variation using the ComBat function in the *sva* R package [26, 28]. Smoking status (current, former, or never-smokers) was estimated from DNA methylation data using previously validated smoking-associated CpG sites [29]. Immune cell-type proportions (CD8, CD4, NK, B cells, monocytes, and granulocytes) were predicted based on the IDOL algorithm [30]. Age, sex, epigenetically inferred smoking status estimated cell proportions, and top 10 within-ancestry PC scores were included as covariates. EWAS were performed separately for each ancestry group. Ancestry-specific results were meta-analyzed using the metagen function available from the meta R package [31]. Both ancestry-specific and cross-ancestry EWAS were used to conduct a differentially methylated-region (DMR) analysis using the dmrff R package [32]. Candidate regions were defined within a 500 bp window with a minimum of two CpG sites. Genomic control correction was applied using the QQperm R package [33]. All association results were corrected for multiple testing using the false discovery rate (FDR q<0.05).

### Summary-based Mendelian Randomization

To assess the effect of genetically regulated DNA methylation changes on GAD and COVID-19 outcomes, we performed summary-based Mendelian randomization (SMR) analysis [34]. We integrated phenotype specific genome-wide association statistics with blood and brain methylation quantitative trait loci (mQTL) datasets (Additional file 1: Table S1). GAD data were derived from a previous GWAS meta-analysis including 97,383 cases and 1,102,617 controls [35]. GWAS data for COVID-19 severe respiratory symptoms (A2; 9,376 cases and 1,776,645 controls), COVID-19 hospitalization (B2; 25,027 cases and 2,836,272 controls), and COVID-19 infection (C2; 125,584 cases and 2,575,347 controls) were obtained from the COVID-19 Host Genetics Initiative (HGI) study [36]. The mQTL reference data included five blood-based [37–39] and two brain-tissue datasets [40], derived from nondissected fetal brain [41] and dorsolateral prefrontal and frontal cortex of adult brain [42, 43]. To further assess fetal-specific effects, we also analyzed the fetal brain mQTL dataset separately [41]. Following developers’ recommendation [34], SMR analysis tested cis-mQTLs within ±2 Mb of each CpG, considering mQTL-p<5×10 and LD pruning (r²= 0.05–0.9). In the heterogeneity test (HEIDI, p > 0.05), up to 20 top cis-SNPs were selected. Reference data were extracted from the European subset of the 1000 Genomes Project (Phase 3). We used METAL [44] to meta-analyze SMR results across tissues and datasets. CpG sites showing no evidence of heterogeneity (HEIDI P > 0.05) were included in the meta-analysis. An inverse-variance weighted fixed-effects model was applied. Between-study heterogeneity was assessed using Cochran’s Q statistic and the I² metric. Bonferroni correction accounting for the number of CpG sites tested was applied to define significant SMR effects.

### Colocalization Analysis

CpG sites showing convergent SMR associations between GAD and COVID-19 were assessed using HyPrColoc [45] to evaluate shared causal variants. A posterior probability (PP) > 0.70 was considered evidence of shared causality. Colocalized CpG signals were further evaluated for their association with gene expression (eQTLs) in blood, brain, and lung tissues.

## Results

After genetic, epigenetic, and phenotypic QC, the final cohort included 893 participants with complete age, sex, ancestry and phenotype information (Additional file 1: Table S2). Sex and age were included as covariates to account for methylation differences, allowing estimation of independent associations. Applying genomic control correction (Additional file 2: Figure S1A and S1B), our cross-ancestry EWAS meta-analysis of GAD identified 162 FDR-significant CpG sites (Figure 1; Additional file 2: Figure S2), with none of them showing significant cross-ancestry heterogeneity (heterogeneity-p>0.2; Additional file 1: Table S3). The most significant GAD association was cg21853055 (beta=0.08, p=1.24X10^-8^). The COVID-19 cross-ancestry EWAS meta-analysis uncovered 151 FDR-significant CpGs (Figure 1) across the three phenotypes tested (Additional file 1: Table S4): moderate symptoms vs controls (CpG N=10, Additional file 2: Figure S3; top-association: cg05557516 beta=0.07, p=1.2X10^-8^), mild symptoms vs controls (CpG N=1, Additional file 2: Figure S4; top-association: cg24009881 beta=0.18, p=1.03X10^-8^), moderate vs mild symptoms (CpG N=140, Additional file 2: Figure S5; top-association: cg21555783 beta=0.05, p=1.14X10^-8^). None of the COVID-19-associated CpG sites showed significant cross-ancestry heterogeneity (heterogeneity-p>0.2; Additional file 1: Table S4). There was no overlap between GAD and COVID-19 at FDR-significant epigenome-wide level. However, two suggestive CpG sites (p<10^-4^) showed convergent associations: cg16944845 mapping to *ZMIZ1* gene was associated with both GAD (beta=0.03, p=1.74X10^-5^) and COVID-19 moderate vs. mild symptoms (beta=0.03, p=1.56X10^-5^) and cg03554028 mapping to *STAB1* gene was associated with both GAD (beta=0.03, p=5.95X10^-5^) and COVID-19 mild symptoms (beta=0.07, p=9.14X10^-5^). No evidence of cross-ancestry heterogeneity was observed for either CpG across GAD and COVID-19 outcomes (I² ≤ 17%, heterogeneity-p>0.29; Figure 2). Among COVID-19 phenotypes, 32 suggestive CpG sites (p<10^-4^) showed convergent associations with consistent effect directions (Additional file 1: Table S5).

**Figure 1.**
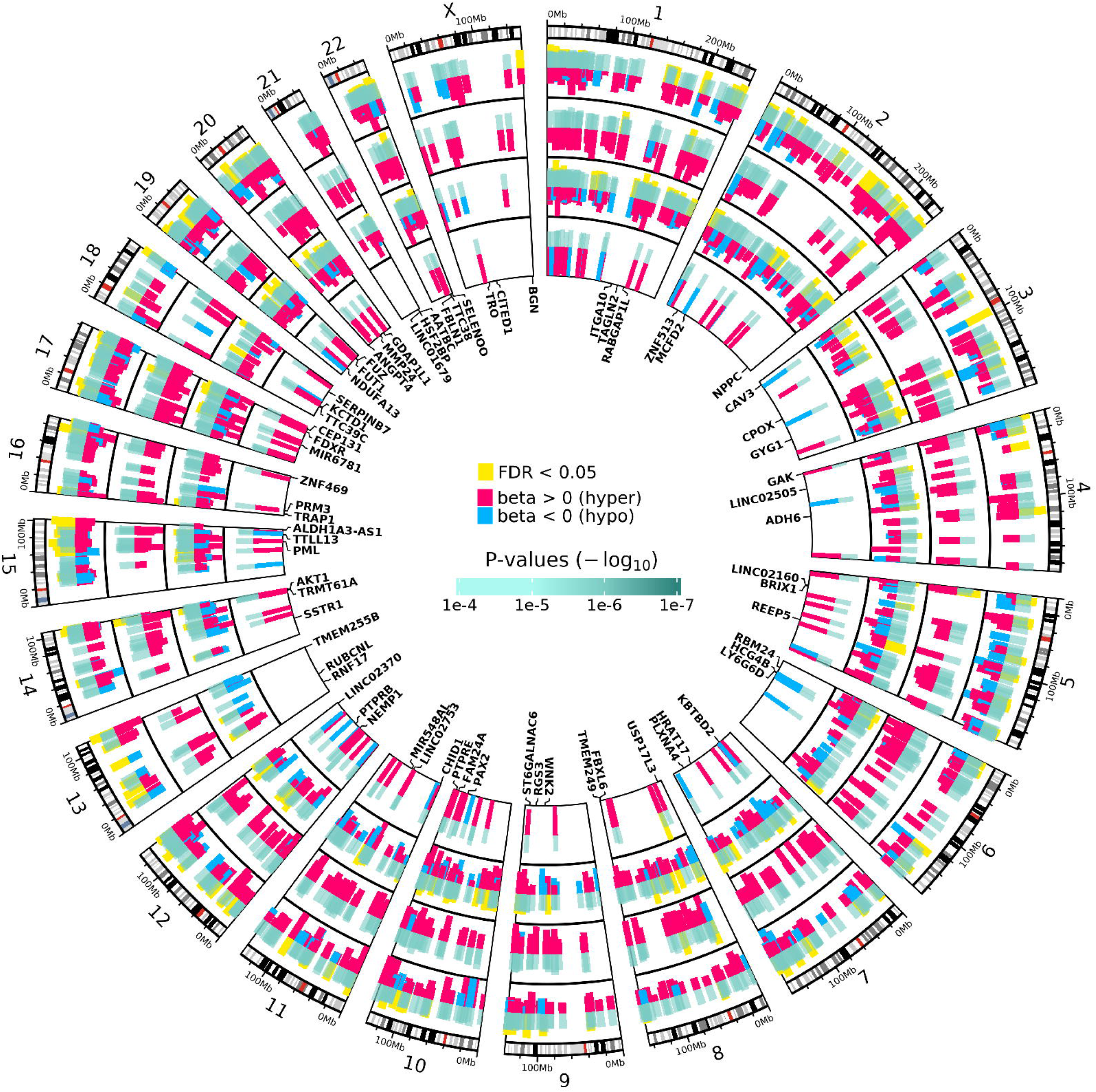
Genome-wide distribution of top EWAS signals for GAD and COVID-19 severity phenotypes. From outer to inner tracks: GAD, COVID-19 moderate vs control, mild vs moderate, and mild vs control. Tracks display the top 200 CpG sites per chromosome (p<1×10□□), with FDR-significant CpGs (FDR q<0.05) highlighted in yellow.

**Figure 2.**
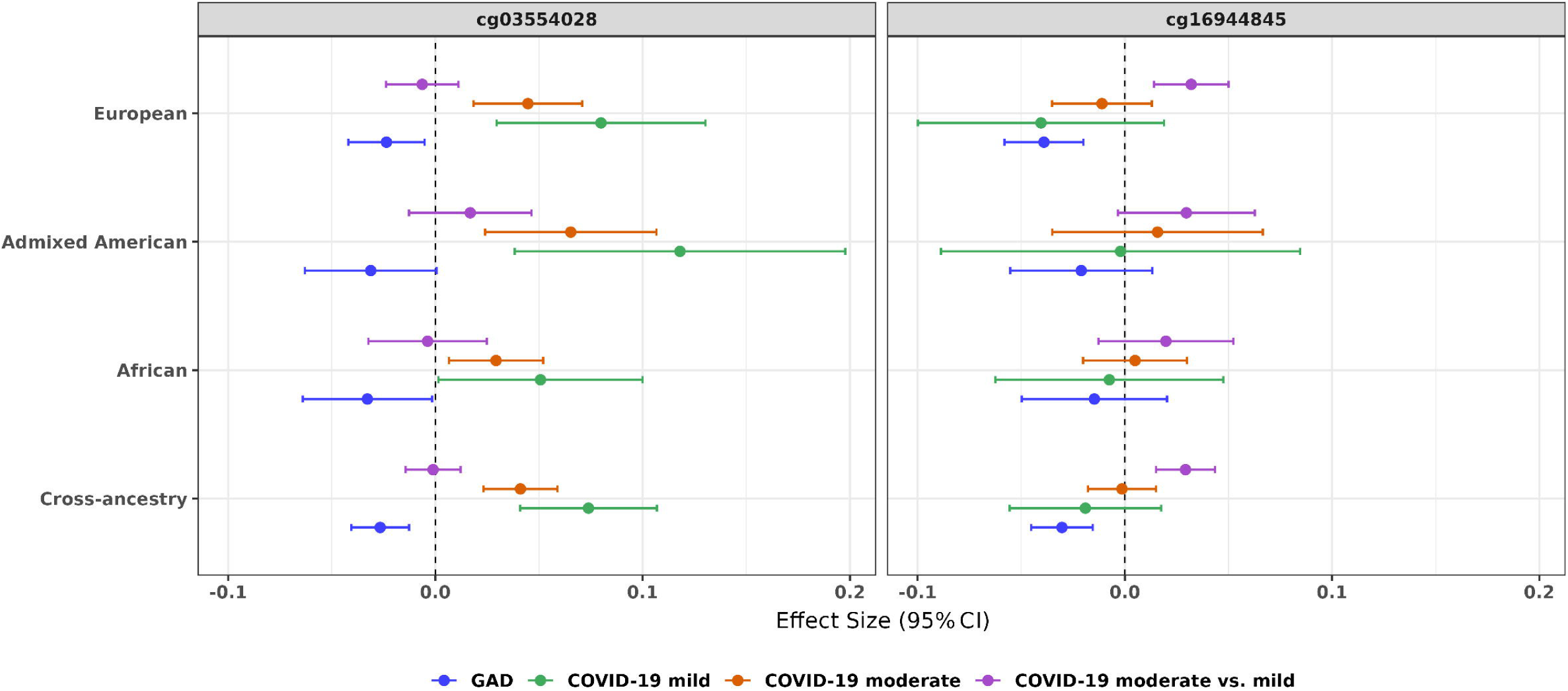
Ancestry-specific and cross-ancestry meta-analytic effect directions of shared CpG sites between GAD and COVID-19 outcomes.

DMR-based analysis allowed us to further expand the epigenetic characterization of GAD and COVID-19 outcomes. A total of 151 FDR-significant DMR-based associations were identified with respect to GAD (Additional file 1: Table S6; Additional file 2: Figure S6). While most GAD-associated DMRs were hypermethylated (∼80%), we identified a tightly clustered hypomethylated region at the *VHL* locus (CpG n=30, length=290 bp, beta= -0.02, p=6.67X10). Among the novel loci, the most significant DMRs (p<10^-10^) included an intergenic region on chromosome 6 (CpG n=13, length=251 bp, beta=-0.097, p=1.30X10^-17^) and a region mapping to *CACNA1A* (CpG n=6, length=160 bp, beta=-0.81, p=1.65X10^-11^), and a region mapping to *TINAGL1* (CpG n=23, length=2,928 bp, beta=0.02, p=1.93X10^-11^).

After FDR multiple testing correction, we also identified 24 DMRs associated with COVID-19 mild symptoms (Additional file 1: Table S7; Additional file 2: Figure S7), 279 DMRs with respect to COVID-19 moderate symptoms (Additional file 1: Table S8; Additional file 2: Figure S8), and 259 DMRs related to COVID-19 “moderate vs mild symptoms” (Additional file 1: Table S9; Additional file 2: Figure S9). Among DMR results not overlapping with EWAS findings, the strongest signals mapped to *PRDM8* (CpG n=12, length=1827 bp, beta=-0.17, p=5.44X10 ¹²) and *PRTN3* (CpG n=7, length=346 bp, beta=0.05, p=1.49X10 ¹²) for mild and moderate symptoms, respectively. The moderate vs. mild contrast further highlighted a large CpG cluster mapped to *ZFP57* (CpG n=24, length=1212 bp, beta=0.096, p=8.81X10^-17^).

Further, we identified 22 FDR-significant DMRs shared between GAD and COVID-19 phenotypes. (Figure 3; Additional file 1: Table S10). The most significant of these mapped to *VWA7* gene (CpG n=19, length=1,146 bp; GAD beta=0.02, p=7.07X10^-10^; COVID-19 moderate beta=0.03, p=8.12X10^-9^; COVID-19 mild beta=0.02, p=1.31X10^-8^). In addition, a strong convergent association was observed at the *BAIAP2-DT* long non-coding RNA locus (CpG n=, length=1,984 bp), showing significant associations with GAD (p=8.43X10^-8^), COVID-19 moderate symptoms (p=5.09X10^-5^), and COVID-19 moderate vs. mild symptoms (p=1.58X10^-11^). Most DMRs showed concordant effect directions among GAD and COVID-19 phenotypes (Additional file 1: Table S10). We also observed two DMRs with discordant effect directions: DMR at *EPS8L1* gene (CpG n=8, length=1,352 bp) was positively associated with GAD (p=7.64X10^-6^) and inversely associated with COVID-19 moderate vs. mild symptoms (p=8.41X10^-5^), and DMR mapping to *STPG2* (CpG n=10, length=802 bp) was positively associated with GAD (p=4.96X10^-8^) and inversely associated with COVID-19 mild symptoms (p=1.08X10^-6^). Additional 31 DMRs showed shared methylation patterns between COVID-19 phenotypes (Additional file 1: Table S10). We also identified DMRs (FDR q<0.05) associated with GAD and COVID-19 outcomes across ancestry groups (Additional file 1: Table S11).

**Figure 3.**
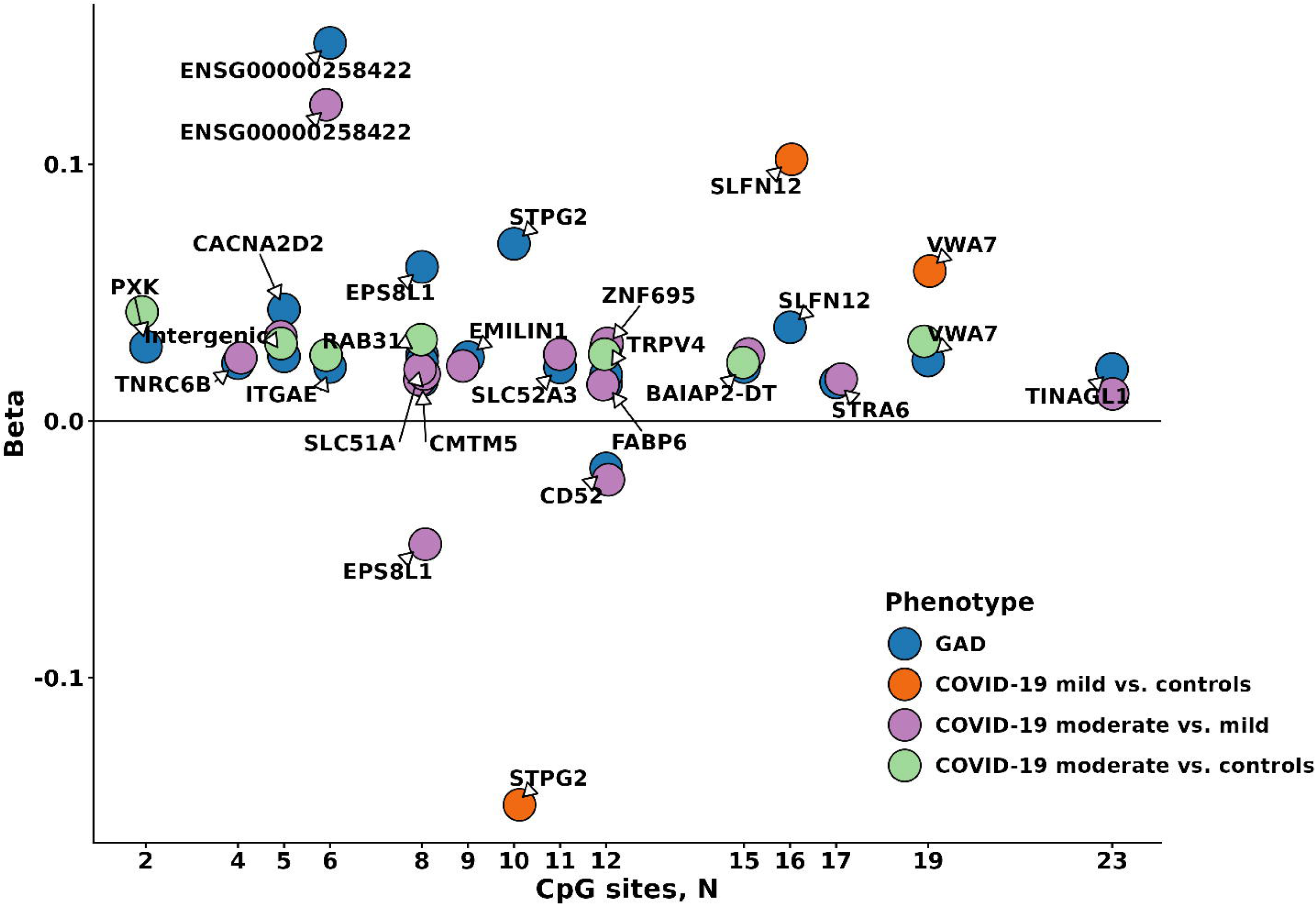
Overlapping differentially methylated regions (DMRs; FDR q<0.05) between generalized anxiety disorder (GAD) and COVID-19 outcomes.

Applying the SMR approach [34] to GAD and COVID-19 HGI genome-wide information [36] and mQTL datasets (Additional file 1: Table S1), we investigated the association of brain and blood genetically regulated methylation changes with GAD and COVID-19 outcomes. In the brain-specific results, multiple CpG sites showed genetically regulated methylation changes associated with GAD (N=15, Additional file 1: Table S12) and COVID-19 phenotypes (infection N=10, Additional file 1: Table S13; hospitalization N=21, Additional file 1: Table S14; severe symptoms N=30, Additional file 1: Table S15). Additional CpG sites were observed when testing mQTL information related to fetal brain (Additional file 1: Table S16). We identified three FDR-significant CpGs (*MICB* cg21619017, *HLA-DPB1* cg02197634, cg02794174) shared between GAD and COVID-19 symptoms in brain-based SMR analysis (Additional file 1: Table S17).

The blood-based SMR meta-analysis identified multiple Bonferroni-significant CpG sites with genetically regulated methylation changes associated with GAD and COVID-19 outcomes (Additional file 1: Table S18-21). Among them, we identified multiple CpG sites (cg00891649, cg05301556, cg07368061, cg09764761) mapping to *MAPT* with evidence of genetically regulated methylation changes positively associated with GAD and inversely associated with COVID-19 severe symptoms and hospitalization (Additional file 1: Table S22). Meta-analyzing SMR estimates obtained from brain and blood mQTL datasets, we further expanded our CpG discovery (Additional file 1: Table S23-26) and identified additional genetically regulated methylation changes overlapping between GAD and COVID-19 (Figure 4; Additional file 1: Table S27). Interestingly, several of the novel CpG sites identified (e.g., *HLA-B* cg01830271, *ZFP57* cg07134666, and *PM20D1* cg26354017) showed the same discordant effect directions (positive association with GAD, inverse association with COVID-19 outcomes) observed with respect to *MAPT*. However, we also observed loci with cross-disorder SMR effects concordant between GAD and COVID-19 outcomes (i.e., cg21657043, cg22103003, *FBXL18* cg00632575, and *STAM2* cg00525834 and cg02156598).

**Figure 4.**
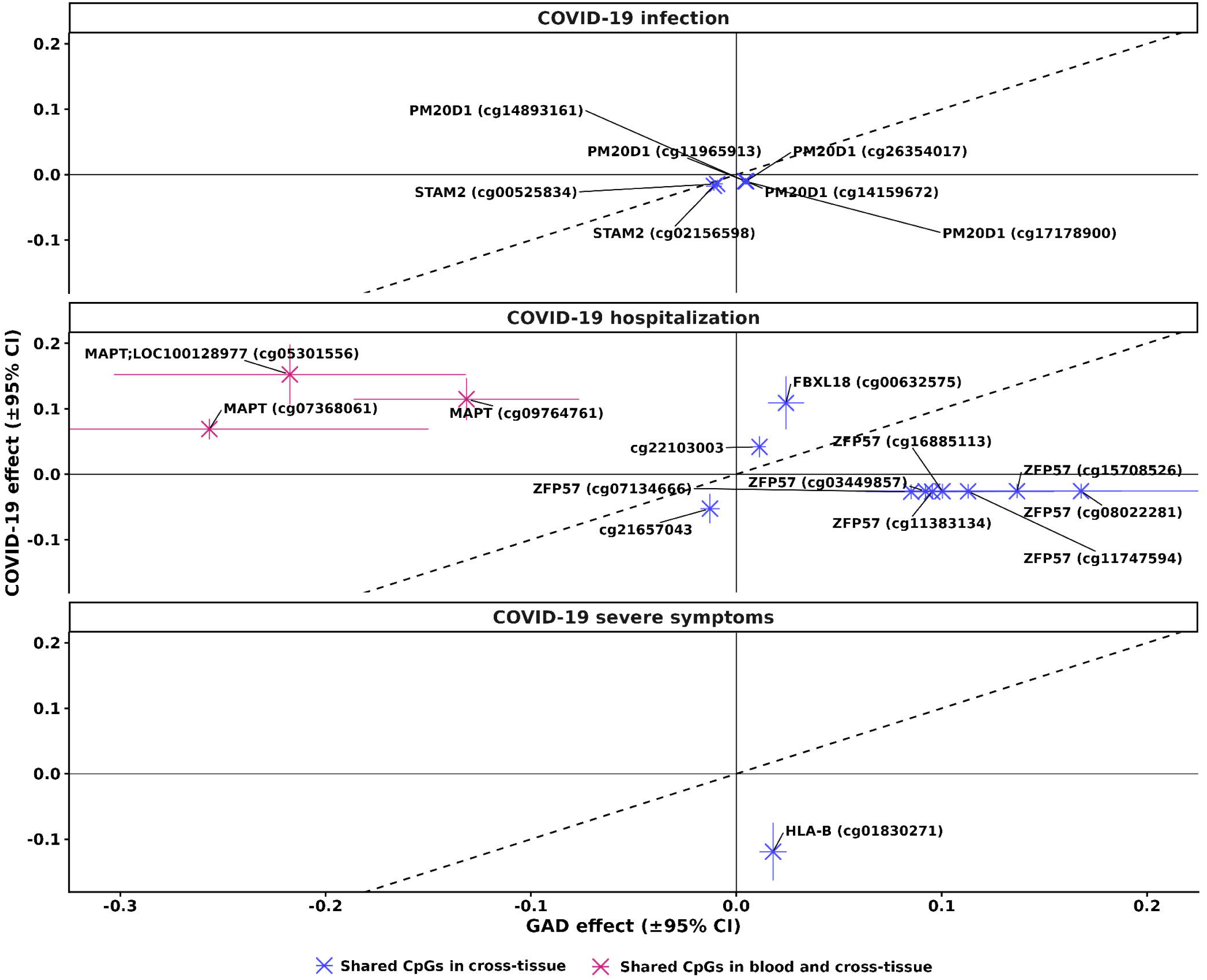
Overlapping CpG loci between GAD and COVID-19 identified in SMR meta-analyses (FDR q<0.05, heterogeneity-p>0.05).

To follow-up SMR results, we conducted a colocalization analysis that identified *FBXL18* rs2966432 as having a potential causal effect on cg00632575 blood-based mQTL, GAD, and COVID-19 hospitalization (PP=0.84). Apart from this shared signal, most colocalized effects linked mQTLs to either GAD or COVID-19, rather than to both phenotypes (Table 1). For instance, rs36092177 showed potential causal effects on GAD and multiple blood methylation signals (N=8, PP>0.7) mapping to *HLA-B* and *ZFP57*. We also identified genetic variants linking COVID-19 phenotypes to multiple CpG sites at the MAPT locus (Figure 5). Among them, rs62056785 effect colocalized with both cg05301556 brain methylation and COVID-19 hospitalization (PP=0.99). Other genetic variants showed colocalized effects linking blood methylation signals to COVID-19 hospitalization and severe symptoms. When testing tissue-specific gene expression, rs2966432 showed colocalized effects with respect to cg00632575 blood methylation, *FSCN1* lung expression, and COVID-19 hospitalization (PP=0.70).

**Figure 5.**
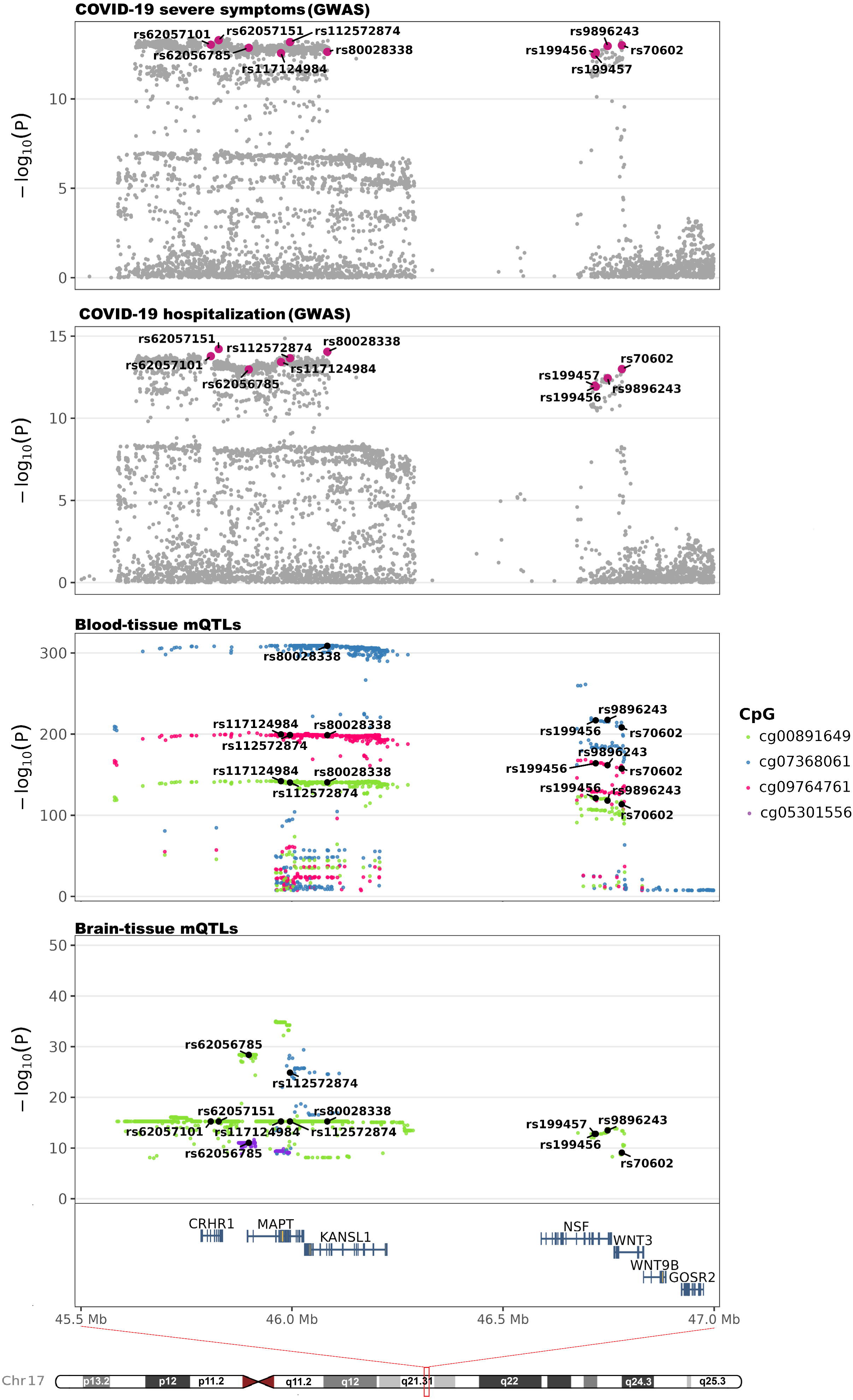
Colocalization between COVID-19 genetic associations and methylation quantitative trait loci (mQTL) in brain and blood within GRCh38/hg38 chr17:45,894,554–46,028,334.

**Table 1:**
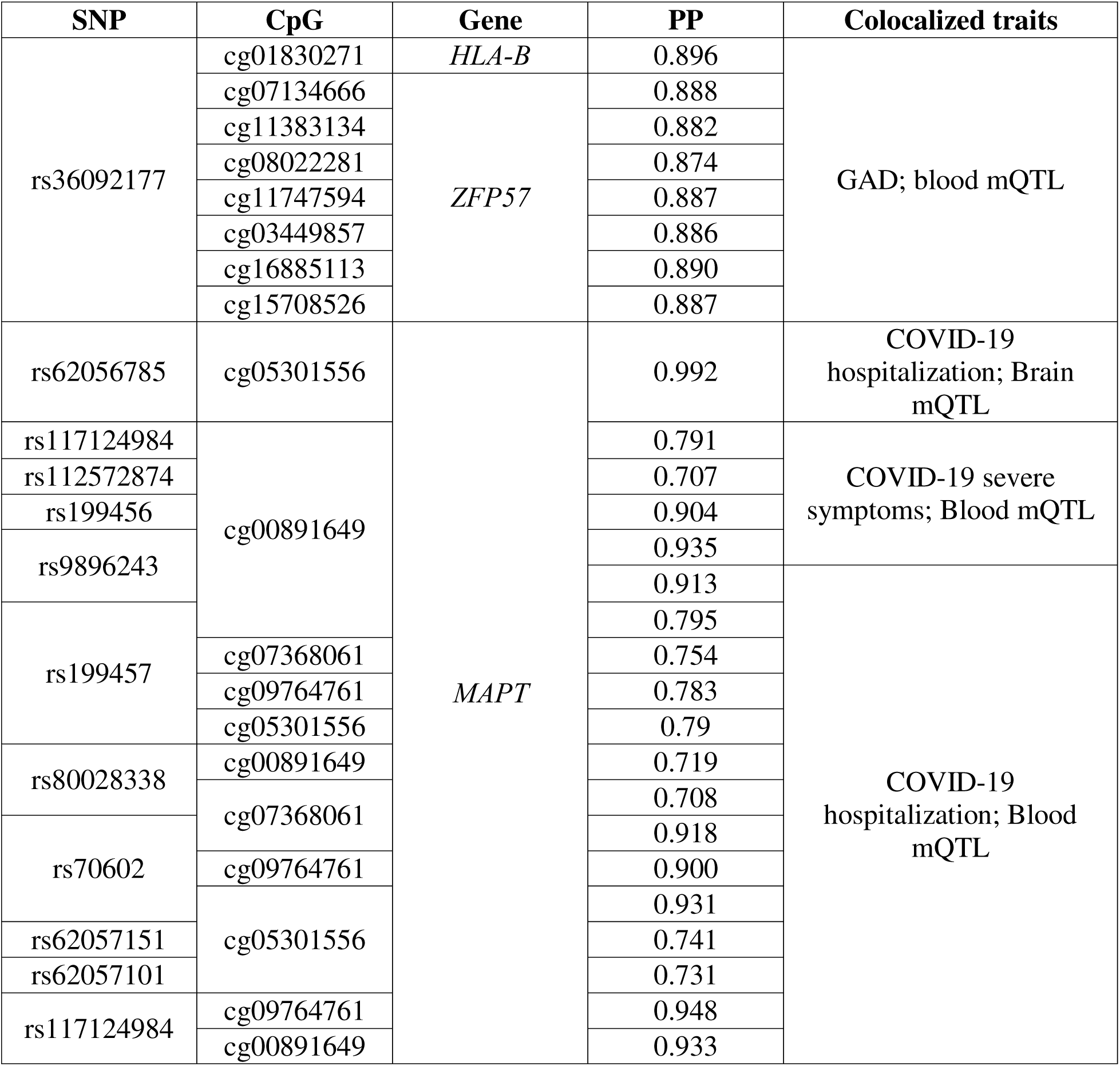
Colocalization of GAD and COVID-19 outcomes with brain and blood mQTLs (posterior probability, PP>0.7).

Assessing the overlap between SMR (genetically regulated methylation changes) and EWAS findings (methylation changes due to the combination of genetic and non-genetic factors), we did not observe any locus surviving strict Bonferroni multiple testing correction. However, considering a suggestive significance threshold (p<10^-4^), we identified loci with convergent EWAS-SMR evidence (Additional file 1: Table S28), which were also supported by a follow-up colocalization analysis (Additional file 1: Table S29). For instance, the methylation of cg01109643 located near *MCF2L* was identified in the blood-based EWAS as being associated with increased GAD (beta=0.04, p=6.49X10^-5^) and in brain SMR as being inversely associated with COVID-19 hospitalization (beta=-0.04, p=3X10^-5^) and severe symptoms (beta=-0.06, p=3.28X10^-6^). With respect to brain-blood convergence, cg08807892 was associated with GAD in the blood-based EWAS (beta=0.07, p=7.36X10^-6^) and in the SMR analysis based both on mQTL datasets derived from brain (beta=0.01, p=7.2X10^-5^) and blood (beta=0.02, p=1.1X10^-5^). The follow-up analysis of these CpG sites showed a colocalization between cg01109643 brain mQTL and COVID-19 hospitalization and severe symptoms (PP=0.92). Additional suggestive evidence for GAD and COVID-19 epigenetic overlap, involving both genetically regulated and non-genetic components, was supported by convergence between DMR and SMR signals across multiple loci (Additional file 1: Table S30 and S31), including *VWA7* and *ZFP57* in blood-based analyses and *MAPT*, *HLA-DPA1*, *FUT1*, *FES*, and *FURIN* in brain-based analyses.

## Discussion

By integrating genome-wide and epigenome-wide data, we conducted a comprehensive study of epigenetic patterns underlying GAD-COVID-19 comorbidity, assessing genetically regulated methylation changes that may link immune and inflammatory pathways to GAD pathogenesis. Results of our cross-ancestry analysis highlighted the consistency of the epigenetic relationship between GAD and COVID-19 across diverse population groups. More broadly, our findings expand the understanding of the methylation underlying the pathogenesis of GAD and COVID-19 and their comorbidity.

Prior epigenome-wide investigations identified loci showing methylation changes associated with anxiety phenotypes, and several of these loci [1, 19, 46] were replicated in our study at epigenome-wide significance (*GALNT9*, *CTBP1*, *LONP1*, *MTA1*, *ZNF574*, *RASGRF2*, *ATP11A*, *PTPRN2*, *SIPA1L3*). Considering the more than 160 GAD-associated CpG sites identified in our study, we observed an enrichment for chromatin regulation, glutamatergic and synaptic signaling, calcium dynamics, cellular stress responses, and immune-related pathways. In particular, GAD-associated CpG sites mapping to *SLC1A3*, *CACNB4*, *CLSTN2*, and *RASGRF2* pointed to a glutamatergic and calcium-dependent synaptic regulatory axis [47–50], which is consistent with the evidence linking synaptic plasticity to anxiety phenotype [51]. Additional loci converged on pathways related to cytoskeletal remodeling and synaptic function. For instance, *AKT1* has been linked to neuronal plasticity in the context of mental illnesses and fear-related brain activation [52, 53], while *LIMK2* function links cytoskeletal remodeling to synaptic function, a pathway broadly involved in neurodegenerative and neuropsychiatric disorders [54]. Several GAD-associated loci (*HDAC4*, *PRDM16*, *CDYL*, and *MTA1*) are implicated in chromatin regulation [55–57]. Notably, *HDAC4* showed strong convergence from both genetic and epigenetic analyses, highlighting its relationship with trauma response, fear behavior, and memory regulation [58, 59]. Another subset of CpG sites mapped to immune-related genes, with *C1QA* pointing to potential involvement in microglia-mediated synaptic remodeling and neuroinflammation [60]. Collectively, these findings suggest that GAD-associated epigenetic changes reflect broader neuroregulatory, stress-related, and immune processes.

GAD-related DMR findings also converged with EWAS signals on loci implicated in glutamatergic, neuronal, and regulatory pathways (e.g., *CACNA1A*, *GRM2*, and *FOXP2*), with this analysis additionally identifying genes involved in extracellular matrix regulation, barrier-vascular homeostasis, metabolic transport, structural remodeling, and immune-inflammatory signaling. Among them, HLA-DPA1/HLA-DPB1 (chr6:33,080,167–33,081,728; 21 CpG sites) supported an association between immune-related methylation and GAD. DMRs mapping to *EGFL7*, *TRPV4*, *EMILIN1*, and *TINAGL1* point to GAD-related methylation changes that may affect barrier–vascular homeostasis, extracellular matrix, and tissue-integrity processes [61–63]. These pathways have been implicated in neuropsychiatric disorders, anxiety and depression-like behaviors, through neurovascular dysfunction, blood–brain barrier disruption, and stress-related remodeling [64, 65].

With respect to COVID-19, several EWAS have been conducted, mostly in relatively small and single-population cohorts [66]. To our knowledge the present study is the largest COVID-19 multi-ancestry epigenome-wide investigation. We replicated loci involved in immune and inflammatory responses (*ZXDC*, *IL17RD*, *PSME3*) [67–69], endothelial and vascular biology (*VEGFB*, *EVL*) [70, 71], and COVID-19 related mortality risk (*CHST8*, *SPATS2L*) [72]. Analyses further revealed several CpG sites mapping to genes not previously highlighted in the COVID-19 epigenetic literature. For example, *ANGPT4* and *PTPRB* are involved in angiogenesis and vascular barrier integrity [73, 74], consistent with evidence linking endothelial injury and dysregulated vascular response to severe COVID-19 [75]. We also identified novel epigenetic evidence linking COVID-19 to immune-related loci such as *GFI1,* which is involved in the proliferation of Th2 cells [76] and *RB1CC1* implicated autophagy-related mechanisms related to SARS-CoV-2 immunity [77]. Particularly notable immune-related locus identified by our EWAS was *CTSB*, given prior evidence of its involvement in SARS-CoV-2 entry process [78]. Our COVID DMR findings identified additional loci involved in neutrophil levels, endothelial dysfunction, cytokine amplification, and prothrombotic vascular injury. DMRs associated with mild COVID-19 mapped to genes involved in endothelial activation (*ANGPT2*) [79], immune signaling (*PIK3CD*) [80], and neutrophil activation (*S100A8*, *PRTN3*) [81, 82]. Conversely, DMRs associated with moderate COVID-19 showed a more pronounced myeloid inflammatory profile, with loci converging on neutrophil degranulation (*ELANE*) [83], chemotaxis (*CXCR2*) [84], myeloid activation (*FCGR2A*) [85], and pro-inflammatory signal amplification (*IRF5*) [86]. Interestingly, DMRs identified in the COVID-19 “moderate vs mild symptoms” analysis highlighted loci linked to platelet-mediated inflammatory processes (*PF4*, *GP1BB*) [87, 88] with broader cytokine-related signaling, including TNF and CSF3 [89, 90]. Our findings align with neutrophil-rich, myeloid-dominant immune dysregulation, endothelial dysfunction, and immunothrombosis reported in COVID-19 [91–93].

To understand possible causal effects linking epigenetic changes to GAD, COVID-19, and their comorbidities, we applied a genetically informed approach that also allowed us to investigate the convergence in methylation regulation between blood and brain. In GAD, our SMR findings pointed to synaptic and neuronal pathways, as well as immune regulatory processes. Among the loci identified, *MAD1L1* was identified in both blood and brain analyses. Prior genetic and methylation studies link this gene to anxiety, PTSD and neurodevelopmental phenotypes [94, 95]. Brain-based SMR analysis uncovered genetically regulated methylation changes mapping to genes involved in brain biology (*NT5DC2*, *FARP1*) and inflammatory processes (*ITIH1*) [96–98]. SMR results observed with respect to fetal brain mQTLs suggested the potential role of mitochondrial biology (*UQCC5*) in GAD pathogenesis [99]. The blood-based GAD SMR meta-analysis uncovered signals related to broad cellular regulatory components (*PRPF40A*, *TAF6*, *WDR46*) [100–102]. Interestingly, SMR meta-analysis integrating both brain and blood mQTLs identified several genes involved in neuronal and synaptic development (*BARHL2*, *FOXP1*, *CTTNBP2*, and *STX1B*) [103–106]. In both blood-only and cross-tissue GAD SMR meta-analyses, we identified a large number of CpGs mapping within an MHC region, which includes genes involved in immune-regulatory components to GABAergic biology (*GABBR1*, *HLA-*G, *HLA-K*, *HLA-J*, *HLA-DPB1*, MICF, C2, *ZFP57*, *HCG11*, *TRIM31*, *TRIM26*, *BAG6*, and *TAPBP*). Given the well-established role of GABAergic signaling in anxiety [107], our findings supported potential interaction between immune-regulatory MHC components and GABAergic pathways, consistent with a neuroimmune interface[108].

With respect to COVID-19, SMR results pointed to host-response mechanisms linked to interferon-related and antigen-presenting pathways. Importantly, SARS-CoV-2 infection was strongly associated with genes involved in respiratory epithelial biology (*ELF5*), previously implicated in a >4-fold higher risk of severe COVID-19 [109], mucosal defense (*MUC1*, *MUC4*) [110], and alveolar epithelial barrier function (*CLDN1*) [111]. However, COVID-19 hospitalization and severe respiratory symptoms were more related to thrombotic processes (*SERPINE1, HLA-DQB1*) [112, 113]. Brain SMR analyses of COVID-19 phenotypes (infection, hospitalization, and severe respiratory symptoms) converged on *XCR1*, *OAS1*, and *OAS3* genes, with supporting evidence also when the analysis was limited to fetal-brain mQTL data. These loci are consistent with the role of antigen-presenting immune biology (*XCR1*) and interferon-mediated antiviral immunity (*OAS1* and *OAS3*) in COVID-19 susceptibility [114]. Brain SMR analyses of COVID-19 hospitalization and severe respiratory symptoms both identified CpG sites mapping to HLA region, a well-established severity locus in COVID-19 biology [115]. Additionally, we observed *FYCO1* locus in COVID-19 SMR analysis based on fetal-brain QTLs. This gene has been implicated in viral intracellular replication and autophagy-related processes [116]. Because this was observed in fetal brain, genetically regulated methylation *FYCO1* changes may be linked to developmental processes implicated into COVID-19 susceptibility.

In EWAS, DMR, and SMR analyses, we observed convergence between GAD and COVID-19 findings. EWAS convergence was limited to two suggestive loci: *ZMIZ1*, associated with neurodevelopmental conditions [117], autoimmune risk [118], and adaptive immune regulation [119]; and *STAB1* has been implicated in inflammatory regulation in atherosclerotic lesions and was identified among shared differentially expressed genes linking COVID-19 and atherosclerosis [120], as well as being implicated in cognitive function [121]. In contrast, 22 DMRs reached FDR significance in both GAD and COVID-19 analyses. Among these, DMRs included genes involved in vascular tissue homeostasis and extracellular matrix biology (*TINAGL1*, *EMILIN1*, *TRPV4*, and *VWA7*). Prior studies highlighted that vascular and blood-brain barrier dysfunction can contribute to COVID-19 related brain changes [122]. In this context, *TRPV4* is especially notable because it has been linked to endothelial-barrier dysfunction [123] and neuroinflammation-associated depression-like phenotypes [124], supporting shared neuroimmune processes linking GAD and COVID-19. Another notable locus was *EPS8L1*, which is related to the *EPS8* pathway that has been implicated in spine remodeling and synaptic plasticity [125]. An additional intriguing pathway is related to retinol. Our DMR analysis identified *STRA6* with respect to both GAD and COVID-19. This gene is involved in cellular retinol homeostasis via RARA-mediated retinoid signaling [126]. A previous independent GAD EWAS identified *RARA* gene [3], which encodes retinoic acid receptor alpha. This locus achieved FDR significance in our COVID-19 EWAS.

Our SMR analysis highlighted 21 CpG sites with genetically regulated methylation changes associated with both GAD and COVID-19. Of these, *FBXL18* cg00632575 showed colocalization among GAD, COVID-19 hospitalization, and blood mQTL. *FBXL18* gene has already been linked to inflammatory processes in response to virus-infected astrocytes [127]. We also observed additional colocalization signals linking mQTLs to disease phenotypes. Specifically, multiple *MAPT* CpG sites showed colocalization of brain and blood mQTLs with both COVID-19 hospitalization and COVID-19 severe symptoms. *MAPT* gene has been linked to both COVID-19 post-acute neurological changes [128] and to stress-related anxiety phenotypes and mood regulation [129]. Blood mQTLs at multiple *ZFP57* CpG sites colocalized with GAD genetic association. This gene is involved in imprinting maintenance and methylation-sensitive chromatin stability that contribute to early factors influencing lifelong health [130]. Testing tissue-specific transcriptional regulation, we uncovered rs2966432 colocalization among cg00632575 blood mQTL, COVID-19 hospitalization, and lung tissue specific *FSCN1* gene expression. This locus plays an important role in dendritic-cell maturation and inflammatory processes [131], and was downregulated in COVID-19 lung biopsies [132]. Among the other genetically regulated methylation associations shared between GAD and COVID-19, *MICB* and *HLA-DPB1* were identified only when testing brain mQTLs. Both loci were linked to stress and infection response and neuroinflammation regulation [133, 134]. This further supports that part of GAD–COVID pleiotropy is driven by epigenetic regulation acting on neuroimmune pathways.

Comparing EWAS/DMR vs. SMR findings allowed us to explore potential differences between observed vs. genetically regulated methylation changes. Indeed, while the former reflects epigenetic modification due to genetic regulation, environmental factors, and the consequences of pathogenic processes, the latter is due only to genetic regulation. Considering brain and cross-tissue analysis, we observed nine COVID-19 SMR findings overlapped with GAD DMRs, mapping to genes involved in adaptive immunity (*HLA-DPA1*) [115], neuroinflammatory changes (*MAPT*) [128], and extracellular matrix remodeling, (*LOXL1*) [135]. With respect to blood and cross-tissue analyses, 17 loci identified by GAD SMR analyses overlapped with COVID-19 DMRs mapping to genes involved in epigenetic regulation and alveologenesis (*ZFP57*) [130], RNA processing (*HNRNPH1*) [136], immune signaling (*FCGR2A*) [137], and metabolic transport biology (*STRA6*, *SLC38A4*) [126, 138]. Among these, *FURIN* is particularly notable because of its role in SARS-CoV-2 infectivity [139], as well as its involvement in neurotrophic and synaptic protein maturation and inflammatory and hypoxic stress responses [140]. We also observed a GAD-COVID-19 convergence between EWAS and SMR findings on *MAPRE3* and *MCF2L* genes, which are involved in cytoskeletal-neuronal remodeling and glutamatergic synaptic organization [141, 142].

While our study expands understanding of epigenetic associations between GAD and COVID-19, we acknowledge several limitations. The cohort investigated in the EWAS and DMR analyses includes participants of diverse ancestral backgrounds. This enabled us to assess possible heterogeneity across population groups, but the sample size available for each ancestral subset may have limited our ability to identify population-specific epigenetic modifications. Similarly, the SMR analyses were limited to populations of European descent, because of the limited sample size of GAD and COVID-19 GWAS and mQTL datasets available for other population groups. Another notable limitation is that only cross-sectional information was available in the cohort investigated in the EWAS and DMR analyses. This did not permit us to investigate how epigenetic changes contribute to the directionality in the comorbidity between GAD and COVID-19. The impact of COVID-19 on anxiety-related outcomes may depend on disease severity. Our severity-stratified analyses addressed this heterogeneity, showing distinct epigenetic patterns across mild and moderate symptoms. However, we could not account for broader contextual stressors associated with infection, including functional impairment, bereavement, socioeconomic or social consequences. In addition, GAD shares genetic liability with other internalizing disorders, but psychiatric comorbidities and pre-existing conditions could not be modeled because of the limited sample size of our cohort. Finally, our analyses were conducted using bulk-tissue epigenetic data, and this may have hidden mechanisms acting on specific cell types.

## Conclusions

In conclusion, this study expands understanding of epigenetic changes associated with GAD and COVID-19, characterizing regulatory mechanisms shared between these conditions. While the GAD methylation profile was primarily enriched for neuronal and synaptic processes, COVID-19 methylation profile was primarily enriched for inflammatory and host-response pathways. Notably, these conditions shared blood and brain epigenetic regulation of genes implicated in stress adaptation, immune activity, and tissue homeostasis. Beyond their relevance to understanding the comorbidity between GAD and COVID-19, these findings highlight the importance of investigating epigenetic changes to elucidate the interplay between mental health and immune function.

## List of abbreviations

CD: cluster of differentiation
CNS: central nervous system
COVID-19: coronavirus disease 2019
CpG: cytosine-phosphate-guanine
DMR: differentially methylated region
EWAS: epigenome-wide association study
FDR: false discovery rate
GAD: generalized anxiety disorder
GAD-7: Generalized Anxiety Disorder 7-item scale
GWAS: genome-wide association study
HEIDI: heterogeneity in dependent instruments
HGI: Host Genetics Initiative
HyPrColoc: Hypothesis Prioritization in multi-trait Colocalization
IDOL: Identifying Optimal Libraries
LD: linkage disequilibrium
mQTL: methylation quantitative trait locus
MWAS: methylome-wide association study
NK: natural killer
PC: principal component
PP: posterior probability
QC: quality control
SMR: summary-data-based Mendelian randomization
SNP: single nucleotide polymorphism

## Additional information

## Supplementary Information

### Additional file 1

Table S1. mQTL datasets tested in SMR.

Table S2. Characteristics of the study cohort.

Table S3. GAD associated CpGs identified in the EWAS analyses.

Table S4. CpGs identified in EWAS analyses of COVID-19 phenotypes.

Table S5. Convergent CpGs across COVID-19 phenotypes identified in EWAS analyses.

Table S6. GAD-associated DMRs.

Table S7. COVID-19 mild symptom–associated DMRs.

Table S8. COVID-19 moderate symptom–associated DMRs.

Table S9. COVID-19 moderate vs. mild symptom–associated DMRs.

Table S10. Convergent DMRs between GAD and COVID-19 and across COVID-19 phenotypes.

Table S11. GAD and COVID-19 associated DMRs in ancestry-stratified analyses

Table S12. GAD associated CpGs in the brain tissue SMR analysis.

Table S13. COVID-19 infection associated CpGs in the brain tissue SMR analysis.

Table S14. COVID-19 hospitalization associated CpGs in the brain tissue SMR analysis.

Table S15. COVID-19 severe symptoms associated CpGs in the brain tissue SMR analysis.

Table S16. CpG sites associated with GAD and COVID-19 in fetal brain SMR analyses.

Table S17. Convergent CpGs across GAD and COVID-19 in blood and brain tissue SMR analyses.

Table S18. CpGs associated with GAD in blood tissue SMR meta-analysis.

Table S19. CpGs associated with COVID-19 infection in blood tissue SMR meta-analysis.

Table S20. CpGs associated with COVID-19 hospitalization in blood tissue SMR meta-analysis.

Table S21. CpGs associated with COVID-19 severe symptoms in blood tissue SMR meta-analysis

Table S22. GAD-COVID-19 convergent signals in blood tissue SMR meta-analysis.

Table S23. CpGs associated with GAD in cross-tissue SMR meta-analysis

Table S24. CpGs associated with COVID-19 infection in cross-tissue SMR meta-analysis

Table S25. CpGs associated with COVID-19 hospitalization in cross-tissue SMR meta-analysis.

Table S26. CpGs associated with COVID-19 severe symptoms in cross-tissue SMR meta-analysis.

Table S27. GAD-COVID-19 convergent signals in cross-tissue SMR meta-analysis.

Table S28. Suggestive evidence of convergence between EWAS and SMR analyses.

Table S29. Colocalization of CpG sites showing EWAS-SMR convergence.

Table S30. GAD DMRs and COVID-19 SMR convergent signals.

Table S31. COVID-19 DMRs and GAD SMR convergent signals.

### Additional file 2

Figure S1A. QQ plots for cross-ancestry and ancestry-specific GAD-EWAS. Figure S1B. QQ plots for cross-ancestry and ancestry-specific COVID-19 EWAS.

Figure S2. Cross-ancestry meta-EWAS results for GAD.

Figure S3. Cross-ancestry meta-EWAS results for COVID-19 moderate symptoms.

Figure S4. Cross-ancestry meta-EWAS results for COVID-19 mild symptoms.

Figure S5. Cross-ancestry meta-EWAS results for COVID-19 moderate vs mild symptoms.

Figure S6. Genome-wide distribution of DMRs for GAD.

Figure S7. Genome-wide distribution of DMRs for COVID-19 mild symptoms.

Figure S8. Genome-wide distribution of DMRs for COVID-19 moderate symptoms.

Figure S9. Genome-wide distribution of DMRs for COVID-19 moderate vs mild symptoms.

## Declarations

### Ethics approval and consent to participate

This study was conducted under protocol #2000033404 approved by the institutional review board of the Yale School of Medicine, New Haven, CT, USA. Written informed consent was obtained from all participants in accordance with the Declaration of Helsinki.

### Consent for publication

Not applicable.

### Availability of data and materials

The PsychGenCOV19 datasets supporting the conclusions of this article are available in the National Institute of Mental Health Data Archive (https://nda.nih.gov/, Collection #4587). Anxiety genome-wide association statistics are available at https://zenodo.org/records/13135834. COVID-19 HGI genome-wide association statistics are available at https://www.covid19hg.org/results/r7/.

### Competing interests

RP and JG are paid for editorial work by the journal Complex Psychiatry. RP received a research grant outside the scope of this study from Alkermes. The remaining authors declare that they have no competing interests. JHK is a scientific advisor to Biohaven Pharmaceuticals, BioXcel Therapeutics, Inc., Cadent Therapeutics (Clinical Advisory Board), PsychoGenics, Inc., Stanley Centre for Psychiatric Research at the Broad Institute of MIT and Harvard, Lohocla Research Corporation. JHK owns stock and/or stock options in Biohaven Pharmaceuticals, Sage Pharmaceuticals, Spring Care, Inc., BlackThorn Therapeutics, Inc., Terran Biosciences, Inc. JHK reports income <$10,000 per year from: AstraZeneca Pharmaceuticals, Biogen, Idec, MA, Biomedisyn Corporation, Bionomics, Limited (Australia), Boehringer Ingelheim International, Concert Pharmaceuticals, Inc., Epiodyne, Inc., Heptares Therapeutics, Limited (UK), Janssen Research & Development, L.E.K. Consulting, Otsuka America Pharmaceutical, Inc., Perception Neuroscience Holdings, Inc. Spring Care, Inc., Sunovion Pharmaceuticals, Inc., Takeda Industries, Taisho Pharmaceutical Co., Ltd. JHK reports income >$10 000 per year from Biological Psychiatry (Editor). JHK received the drug, Saracatinib from AstraZeneca and Mavoglurant from Novartis for research related to NIAAA grant ‘Centre for Translational Neuroscience of Alcoholism [CTNA-4]’ from AstraZeneca Pharmaceuticals. JHK holds the following patents: 1) Seibyl JP, Krystal JH, Charney DS. Dopamine and noradrenergic reuptake inhibitors in treatment of schizophrenia. US Patent #:5447948. September 5, 1995; 2) Vladimir, Coric, Krystal, John H, Sanacora, Gerard – Glutamate Modulating Agents in the Treatment of Mental Disorders US Patent No. 8778979 B2 Patent Issue Date: July 15, 2014. US Patent Application No. 15/695164: Filing Date: 09/05/2017; 3) Charney D, Krystal JH, Manji H, Matthew S, Zarate C, – Intranasal Administration of Ketamine to Treat Depression United States Application No. 14/197767 filed on March 5, 2014; United States application or Patent Cooperation Treaty (PCT) International application No. 14/306382 filed on June 17, 2014; 4): Zarate, C, Charney, DS, Manji, HK, Mathew, Sanjay J, Krystal, JH, Department of Veterans Affairs ‘Methods for Treating Suicidal Ideation’, Patent Application No. 14/197.767 filed on March 5, 2014 by Yale University Office of Cooperative Research; 5) Arias A, Petrakis I, Krystal JH. – Composition and methods to treat addiction. Provisional Use Patent Application no. 61/973/961. April 2, 2014. Filed by Yale University Office of Cooperative Research.; 6) Chekroud, A., Gueorguieva, R., Krystal, J.H. ‘Treatment Selection for Major Depressive Disorder’ [filing date June 3, 2016, USPTO docket number Y0087.70116US00]. Provisional patent submission by Yale University; 7) Gihyun, Yoon, Petrakis I., Krystal J.H. – Compounds, Compositions and Methods for Treating or Preventing Depression and Other Diseases. U.S. Provisional Patent Application No. 62/444552, filed on January 10, 2017 by Yale University Office of Cooperative Research OCR 7088 US01; and 8) Abdallah, C., Krystal, J.H., Duman, R., Sanacora, G. Combination Therapy for Treating or Preventing Depression or Other Mood Diseases. U.S. Provisional Patent Application No. 62/719935 filed on August 20, 2018 by Yale University Office of Cooperative Research OCR 7451 US01. All other authors report no conflicts.

### Funding

This study was funded by the National Institute of Mental Health (RF1MH132337). SK acknowledges financial support from The Scientific and Technological Research Council of Türkiye (TÜBİTAK) under the 2219 International Postdoctoral Research Fellowship Program. BCM acknowledges support from the MQ Transforming Mental Health (UFA21\100014). JH acknowledges support from the American Foundation for Suicide Prevention (PDF-0-065-23).

### Authors’ contributions

SK and RP designed the study. SK, BCM, JH, DQ, and DD performed or contributed to the analyses. YZN, JHK, RHP, and JG contributed to the phenotyping of the PsychGenCOV19 cohort. AML and JG performed or supported the laboratory experiments performed on the PsychGenCOV19 samples. All the authors participated in the interpretation of data and critical revision of the manuscript for important intellectual content. RP obtained the primary funding. RP supervised the study. All authors read and approved the final manuscript.

## Acknowledgments

The authors thank PsychGenCOV19 participants and acknowledge the important work done by the personnel refining the assessment and involved in the participants’ enrollment.

