## Supplementary Figures for "Shared epigenetic regulation acting on neuroimmune pathways contributes to the comorbidity between generalized anxiety disorder and COVID-19"

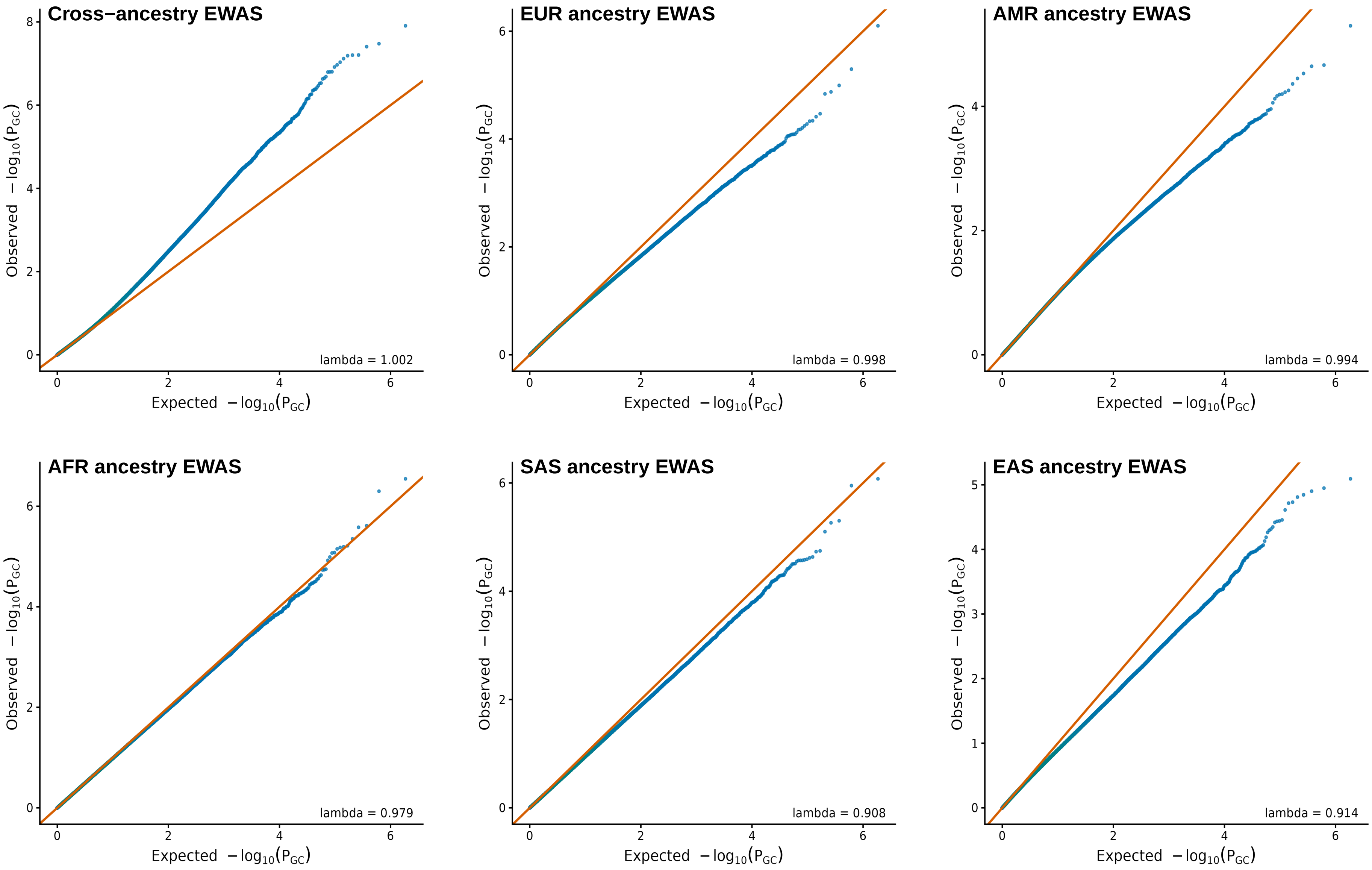


**Figure S1A.** QQ plots based on GAD phenotype cross−ancestry meta EWAS and EWAS in ancestry groups.

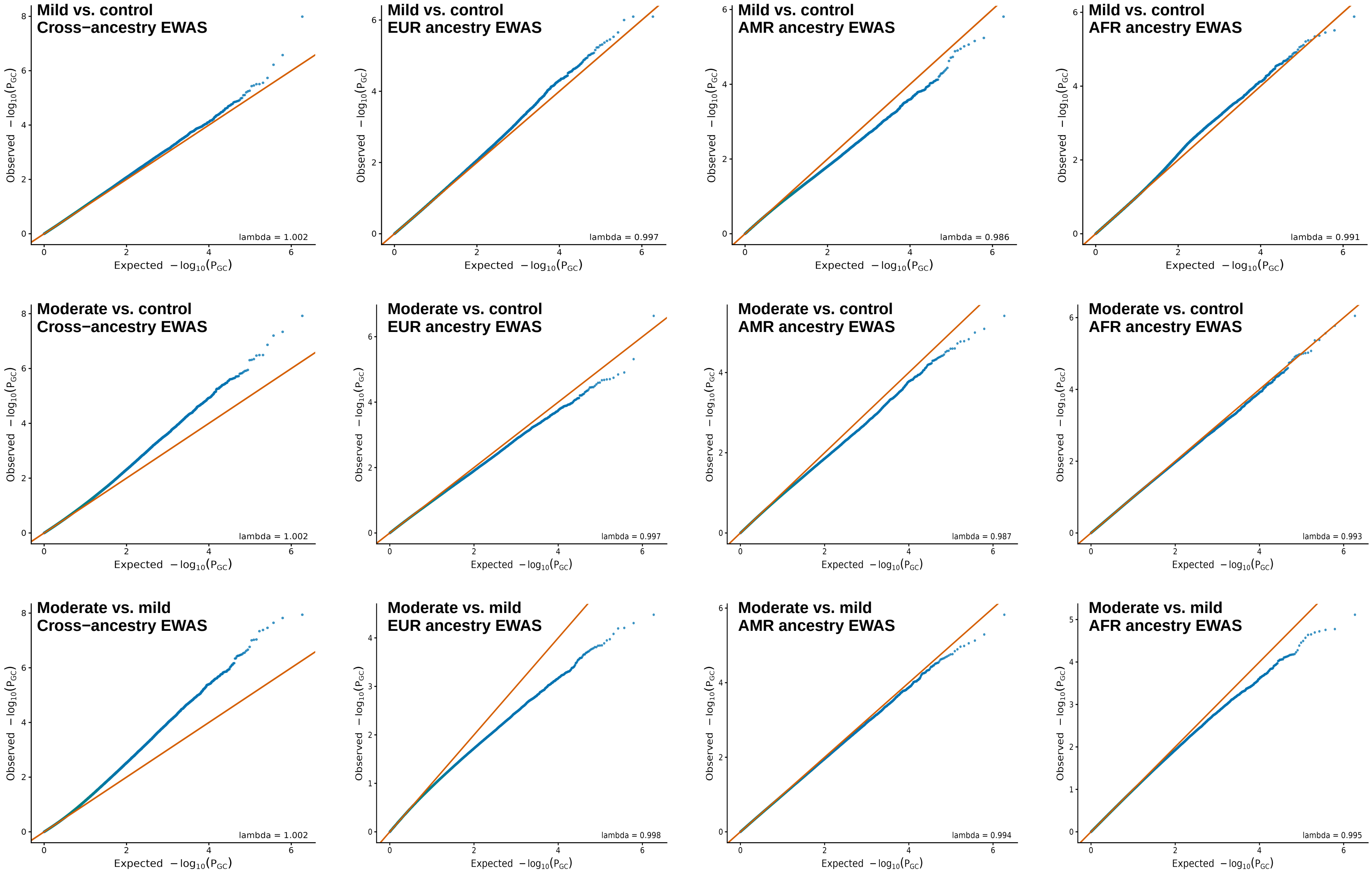


**Figure S1B.** QQ plots based on COVID−19 phenotypes cross−ancestry meta EWAS and EWAS in ancestry groups.

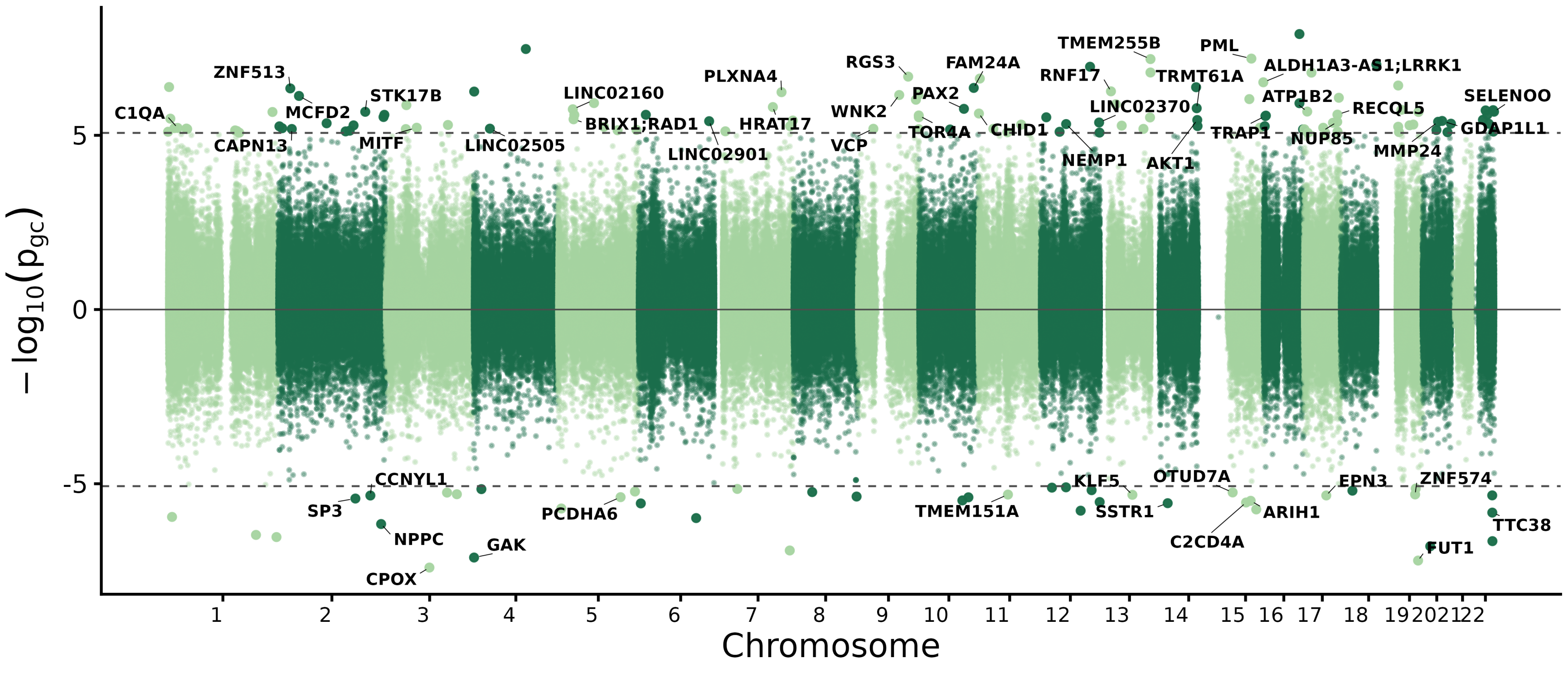

**Figure S2.**Methylation signals associated with GAD phenotype in a cross−ancestry meta EWAS analyses (significance threshold: FDR < 0.05).

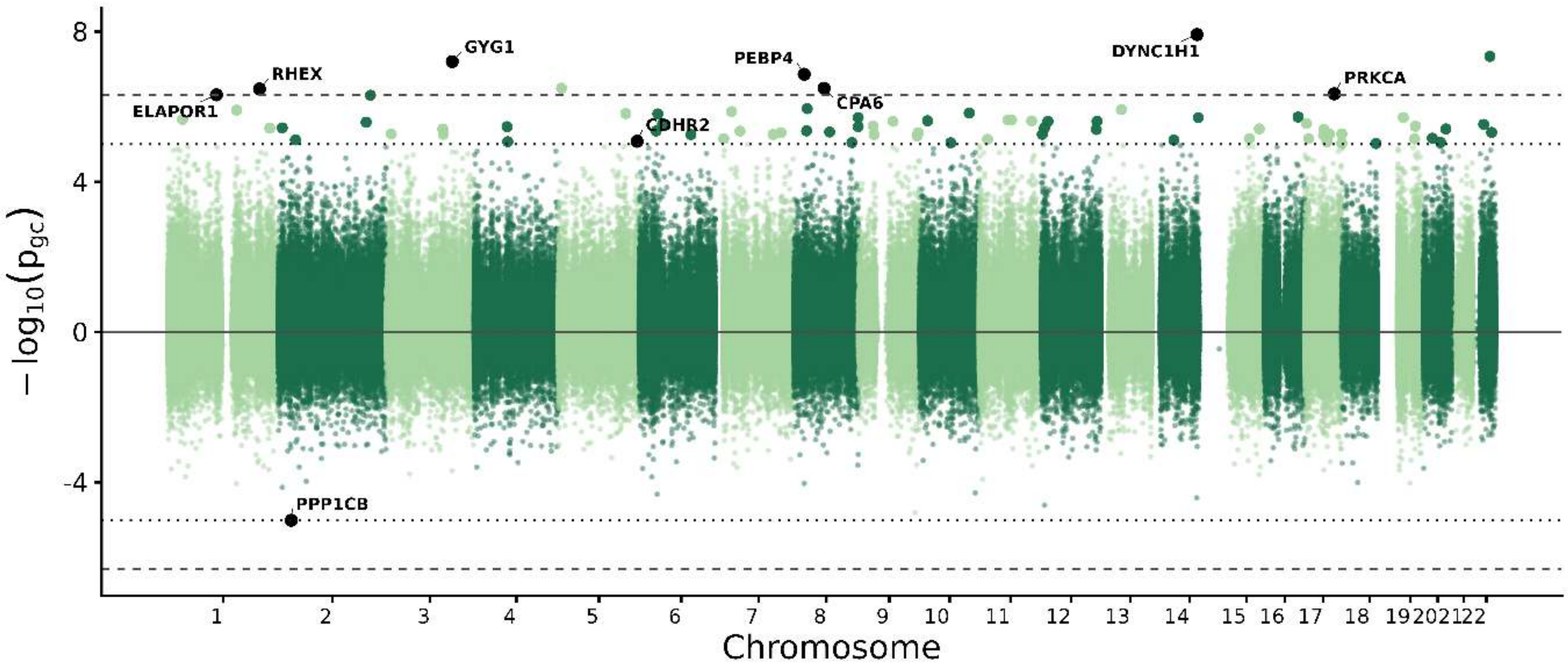


**Figure S3.**Methylation signals associated with COVID−19 moderate symptoms vs. controls in a cross−ancestry meta−EWAS analyses (significance threshold: FDR < 0.05).

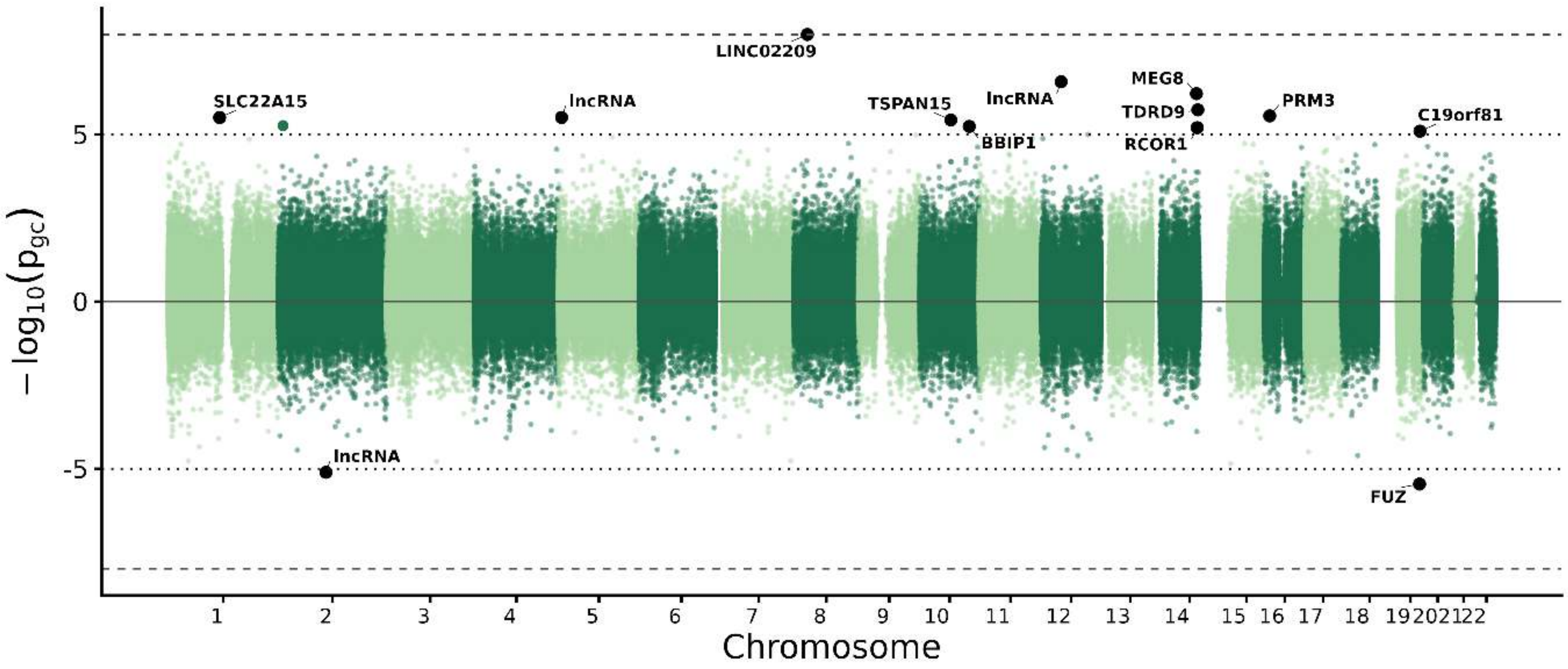


**Figure S4.** Methylation signals associated with COVID−19 mild symptoms vs. controls in a cross−ancestry meta−EWAS analyses (significance threshold: FDR < 0.05; $P<1\times10^{-5}$).

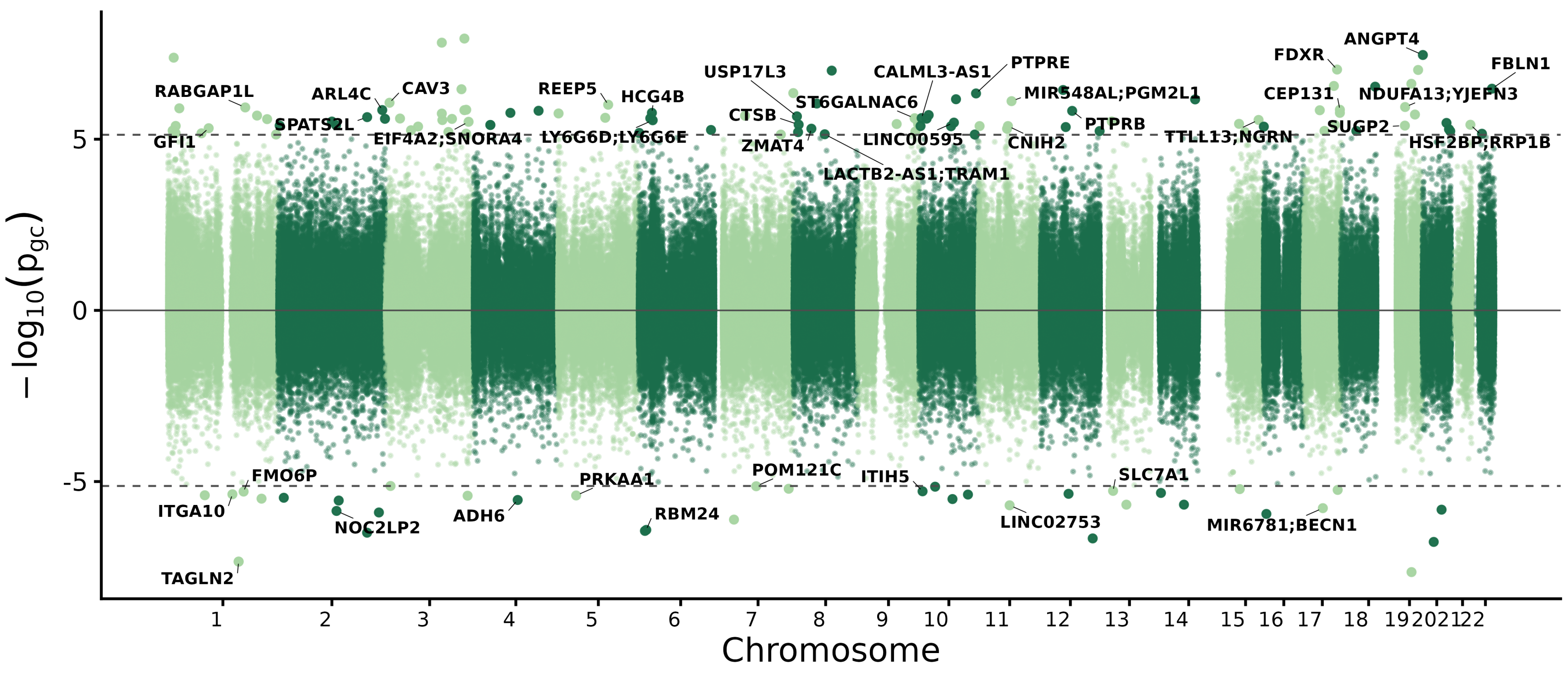


**Figure S5.** Methylation signals associated with COVID−19 moderate vs mild symptoms in a cross−ancestry meta−EWAS analyses (significance threshold: FDR < 0.05).

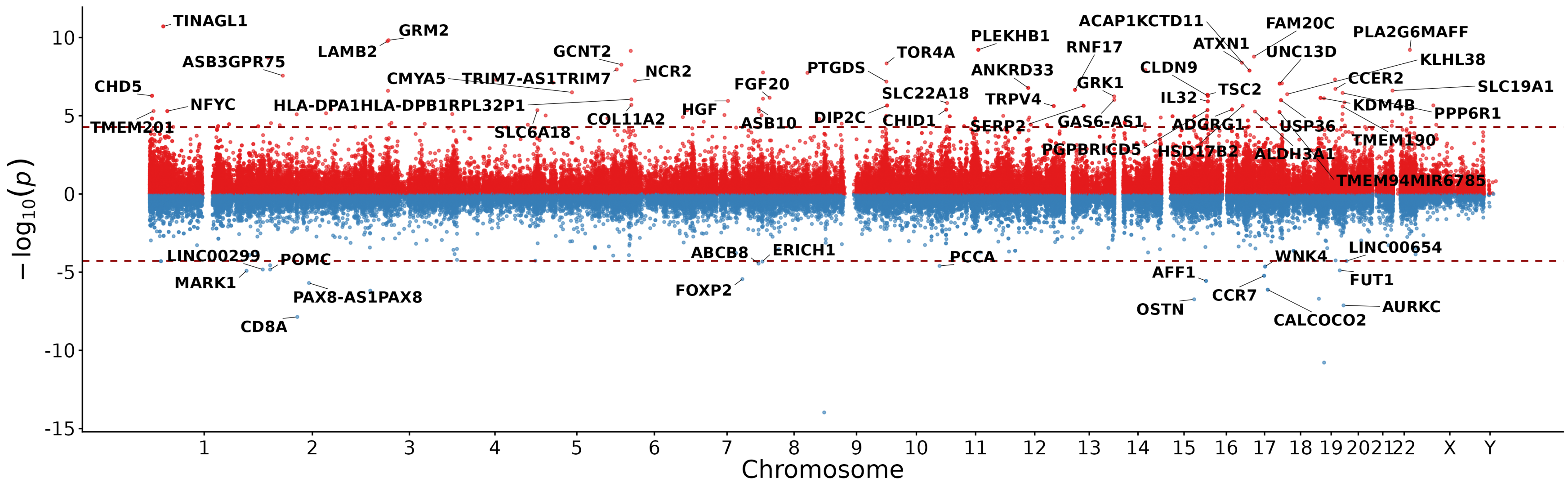


**Figure S6.** Genome wide distribution of GAD associated differentially methylated regions (significance threshold: FDR < 0.05).

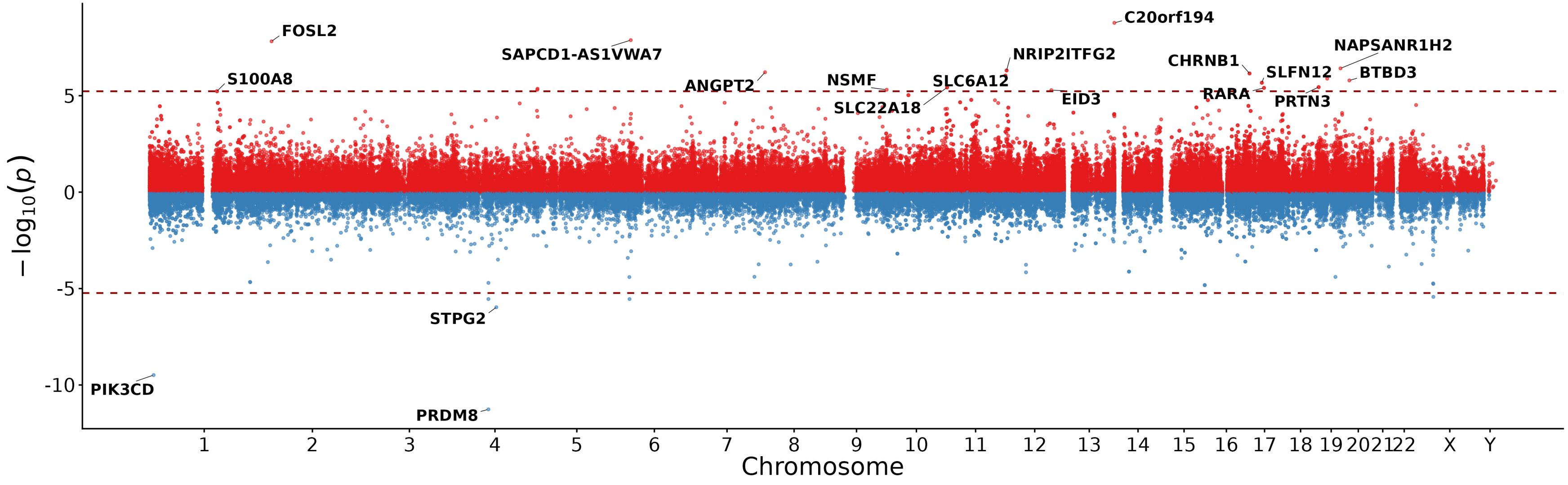


**Figure S7.** Genome wide distribution of COVID−19 mild symptoms associated differentially methylated regions (significance threshold: FDR < 0.05).

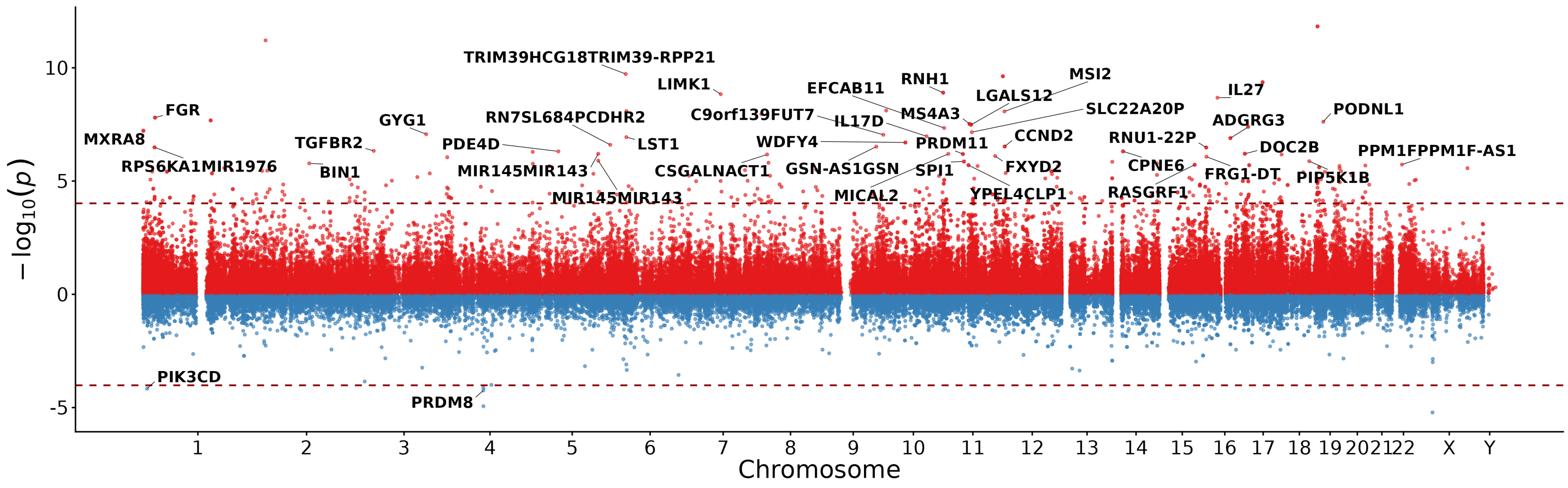


**Figure S8.** Genome wide distribution of COVID−19 moderate symptoms associated differentially methylated regions (significance threshold: FDR < 0.05).

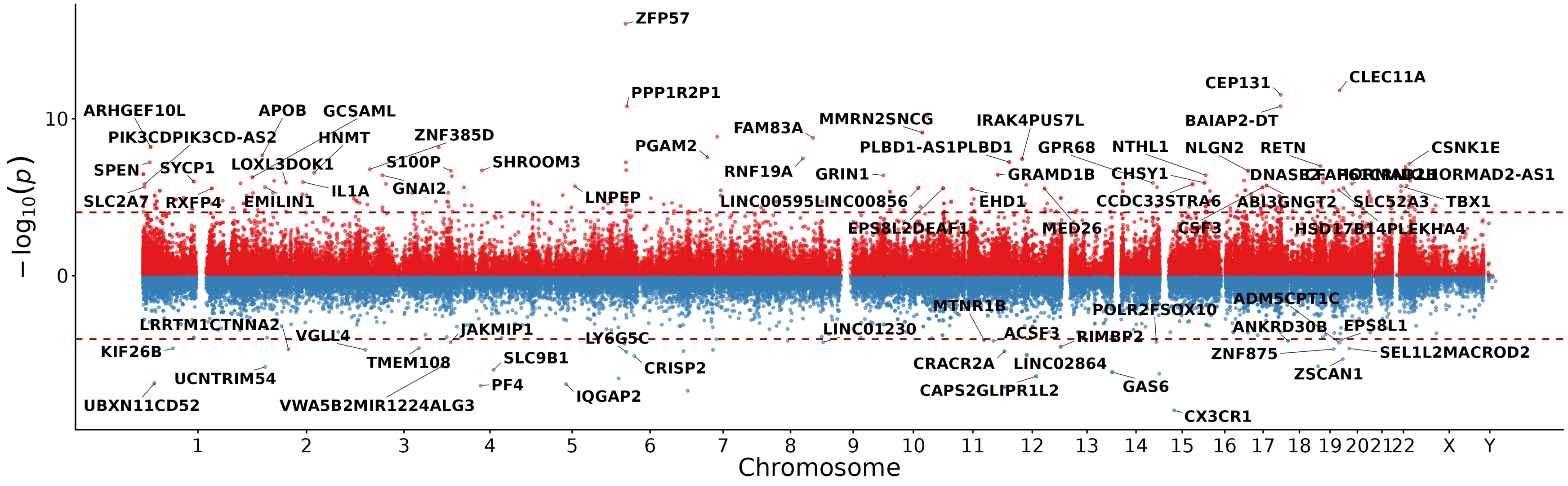


**Figure S9.** Genome wide distribution of COVID−19 moderate vs mild symptoms associated differentially methylated regions (significance threshold: FDR < 0.05).
